# A retrospective analysis to compare the dosimetric parameters of two rectum-sparing techniques during intracavitary brachytherapy with tandem-ring applicators for cervical cancer

**DOI:** 10.1101/2024.12.06.24318598

**Authors:** Siddharth Vats, Shabnum Thakur, Manish Gupta, Vineet Kumar, Lalit Chandrakant, Muninder Negi, Parul Sharma, Vikas Fotedar, Purnima Thakur, Deepak Tuli, Poorva Vias, Jyoti Sharma, Anup Negi, Shilpa Kaushal, Mrinalini Upadhyay, Naina Negi, Ajay Sharma, Anjali Azad, Neeraj Parihar, Divyajyoti Verma

## Abstract

**Purpose:** To compare the standard International Commission on Radiation Units and Measurements-38 (ICRU-38) rectum and bladder point doses with a modified technique of combining radio-opaque vaginal packing (VP) and rectal retractor (RR) versus the conventional technique of using RR alone for rectum separation during high-dose-rate (HDR)-intracavitary brachytherapy (ICB) with tandem-ring (TR) applicators.

**Methods:** Between January 2021 and August 2022, 41 cancer patients received at least one ICB session with the conventional and at least one with the modified technique using the same applicator set and loading pattern. The remaining ICB sessions, were done using that technique, which gave the best dosimetric parameters during previous sessions in that particular patient.

**Results:** Out of total 111 ICB sessions with TR applicators, 44 constitute conventional group and 67 modified group. For the same dose (100%) prescribed to point A in both groups, the mean dose to the ICRU rectal point was 44.1% of the prescription dose (range:23.8-77.8%, median 42.5%) in modified group and 55.5% (range: 36.4-73.1%, median 56.5%) in the conventional group. There was 11.4% reduction in mean dose to the ICRU rectal point (p<0.001). The mean dose to the ICRU bladder point was 55.5% of the prescription dose (range, 14.8-127.2%; median 55.5%) in the modified group and 49.8% (range 11.6-95.6%; median 51.5%) in the conventional group. There was 5.7% increase in the mean dose to the ICRU bladder point (p=0.21). The other point doses and volume parameters were similar between groups.

**Conclusion:** Modified technique of combining rectal retractor with customised radio-opaque vaginal packing, significantly reduced the ICRU rectal point dose, compared to using a rectal retractor alone during brachytherapy with tandem-ring applicators.

## Introduction

The external beam radiotherapy (EBRT) with concomitant chemotherapy combined with intracavitary brachytherapy (ICB) is the standard treatment for patients with locally advanced cervical cancer (LACC)). EBRT and ICB are combined to the total dose of 80 to 90 Gy, traditionally prescribed to point A.^1,2^ EBRT to whole pelvis to the dose of 45-50.4Gy @ 1.8-2Gy per fraction is delivered, which regresses the involved parametrium and tumor shrinks to the extent that now it can be very well encompassed within the high dose region of ICB.^1,3^ This favorable tumor response to EBRT not only makes the patients eligible for ICB but is also associated with spectrum of changes in the normal anatomy at the time of ICB.^4–6^ At one end of the spectrum, there are patients with roomy vagina, and at the other end, there is a subset of patients with narrow vagina and shallower fornices.^6^ During ICB, the potential of rectum toxicity is a significant concern. This is because the vaginal walls have a natural inclination to collapse and remain in close proximity, thereby drawing nearby organs such as the rectum closer to the radiation sources housed within the applicators. Concerted efforts are being made to minimize rectum exposure by integrating sophisticated imaging techniques or by enhancing the techniques aimed at increasing the distance between the rectum and the applicators. The rectal retractor (RR) is the predominant method used to separate the rectum from tandem-ring (TR) applicators during ICB. Literature has indicated that tandem-ring (TR) applicators with rectal retractor (RR) are ideal for patients characterized by shallow fornices and a narrow vagina.^7–9^ Nevertheless, the current trend indicates that their utilization is not limited to the aforementioned subset; rather, it encompasses all individuals undergoing ICB, regardless of their vaginal capacity. In the case of patients with diverse vaginal capacities, the pre-fabricated set of standard RR blades provided by the manufacturer is akin to a one-size-fits-all solution.

A limitation during brachytherapy implant procedure is that during conventional ICB procedure with TR applicators, when RR blade is assembled, the remaining space in posterior fornices becomes inaccessible for packing (Figure 1). This modified technique enables us to overcome this limitation by making the customised vaginal packing (VP) possible for a range of spaces, posterior and superior to RR, in different patients. It helps to supplement the displacement achieved with RR alone, which may lead to reduction in the dose to the ICRU rectal point (D_ICRU_) (Figure 2). A clear correlation between late rectal morbidity (proctitis, bleeding, and fistula) and dose point (D_ICRU_) and dose-volume (D_2cc_, D_1cc_ and D_0.1cc_ i.e. minimum dose to most exposed 2cc, 1cc and 0.1cc volumes of rectum) has already been established in literature.^10–12^ In our previous study, the average reduction in the mean dose to ICRU rectal point was 13% (p=0.001) per ICB session with modified technique compared to conventional approach of using RR alone.^6^ Study had limitations, as the majority of patients in two comparison groups were different. The individual anatomical variation among patients could have been a possible confounding factor and possibility of bias in selecting patients with favourable anatomy for a particular method of rectum separation could not be ruled out. Now this larger study was planned with the aim to see whether similar results are achieved when both methods are applied on the same patient to eliminate confounding factor of individual anatomical variation and to address the bias of selecting a patient for a particular method of rectum separation. Other possible variables related to the physical parameters of applicator were also addressed. We aim to describe in sufficient detail the steps taken during ICB procedure, to enable centres across the world, to practice and evaluate this technique, irrespective of whether they are planning 2-D brachytherapy or 3-D image guided brachytherapy (IGBT). The results of this study in poster format were presented in annual European Society for Radiotherapy and Oncology (ESTRO) congress-2023 at Vienna.^13^

**Figure 1.**
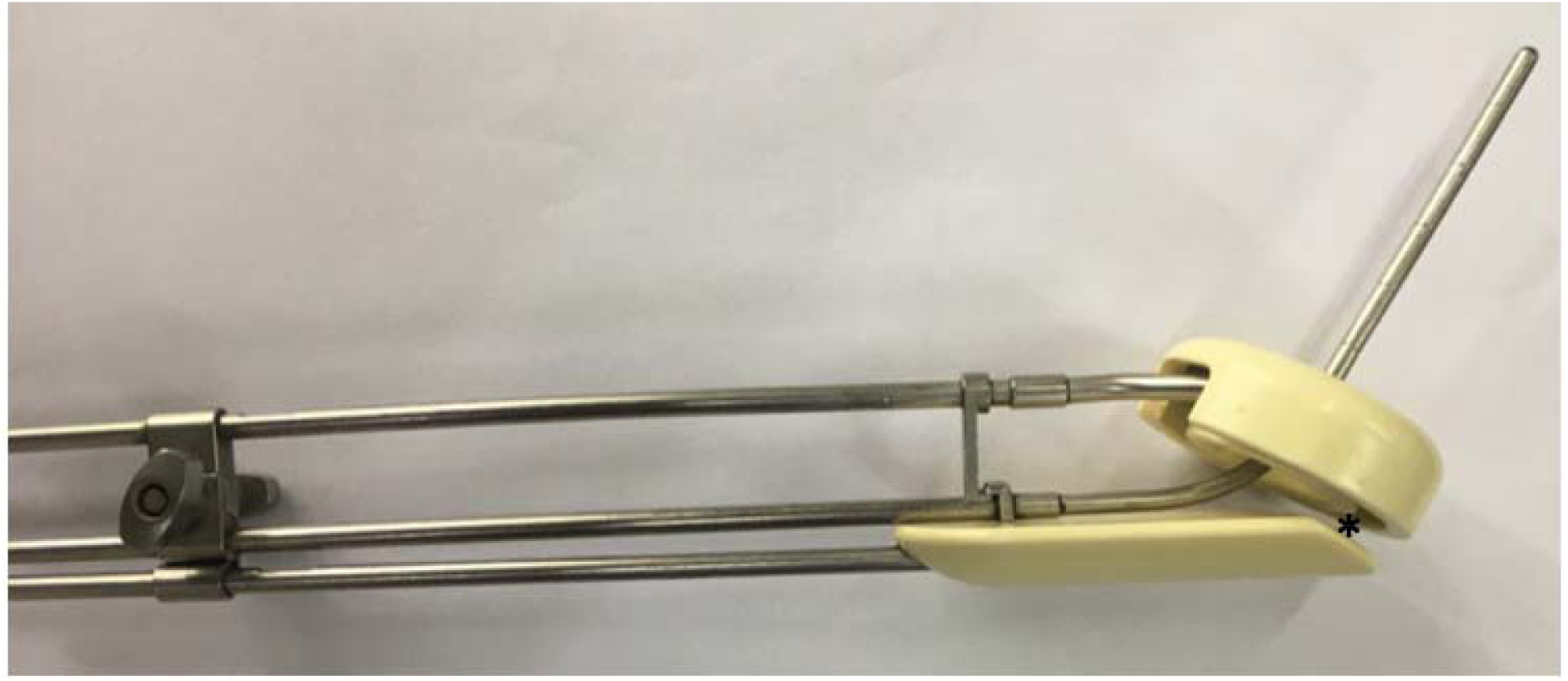
Nucletron tandem-ring(TR) applicator and a rectal retractor (RR) with a tandem length of 6cm, angle of 30^0^ and a ring size of 3cm. (The design best conforms to the anatomy of the patients in ideal subset described in the text, but in other patients, especially with roomy vagina, due to an inadequate gap (*) as the sloping cranial edge of RR closely abuts the posterior aspect of the ring, the potential space posterior and superior to the cranial edge of RR becomes inaccessible for vaginal gauze packing. The modified technique enabled us to overcome this limitation and allowed customised vaginal gauze packing for optimal displacement of the rectum).

**Figure 2.**
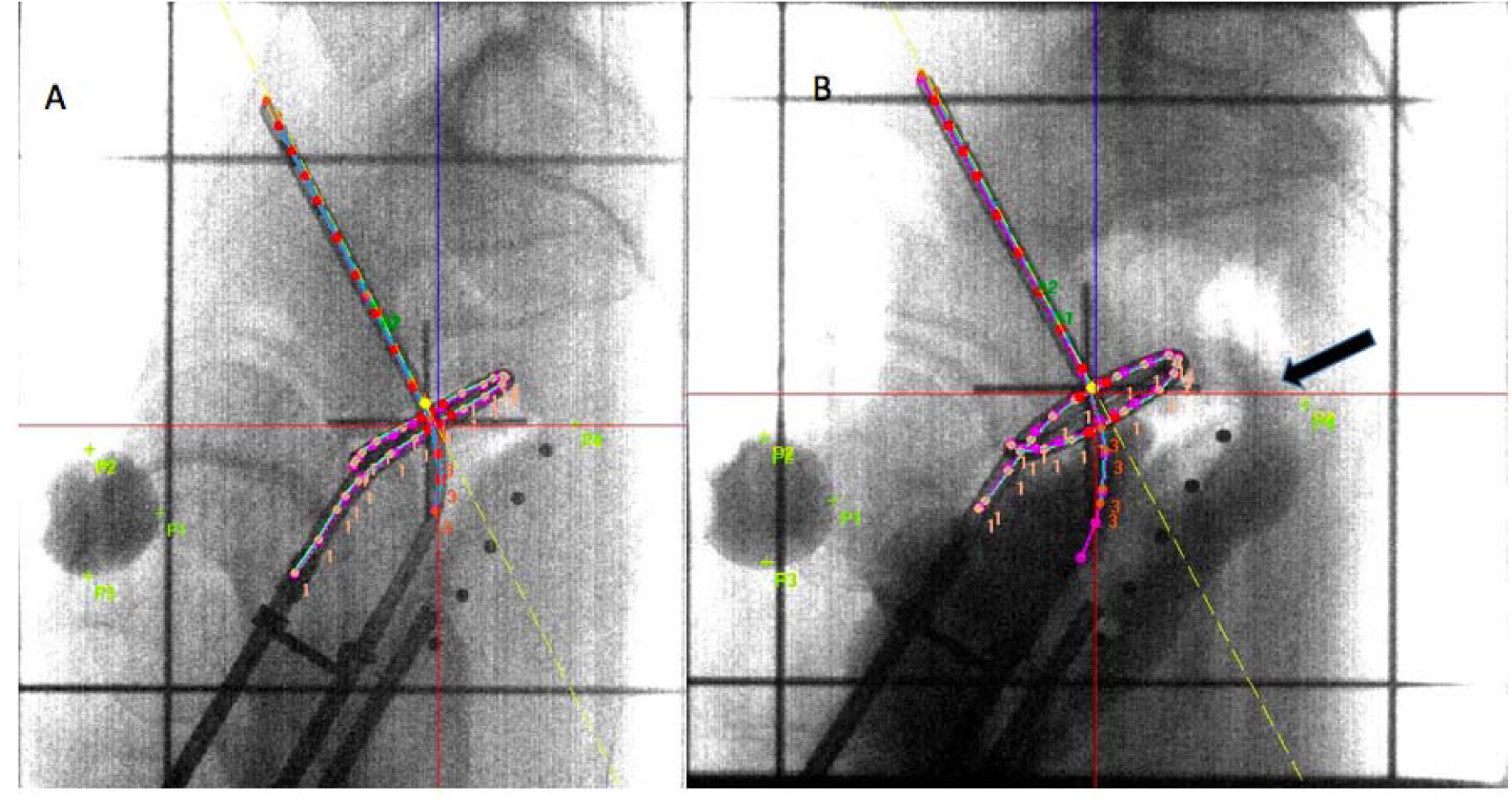
(A) Simulation image of applicators (shown in Figure 1), with conventional technique of using rectal retractor alone. (B) Modified technique of combining radio-opaque vaginal gauze packing and rectal retractor (↑) applied using same applicator set and loading pattern (without optimization), in the consecutive session on one of the patients. The modified technique resulted in a reduction of 1.1 Gray (16%) in the ICRU rectal point dose, while delivering 7 Gray (100%) to point A in both sessions. ICRU: International Commission on Radiation Units and measurements; P_1_: ICRU bladder point; P_4_: ICRU rectal point.

## Materials and methods

The records and treatment plans of locally advanced cervical cancer (LACC) patients with stage IB2 to IVA as per International Federation of Gynaecology and Obstetrics-2018 (FIGO-2018), who received treatment with curative intent, between January 2021 to August 2022, were studied. This time frame was chosen due to the continuous improvements implemented previously, which resulted in the development of a standardized protocol in our department for the management of LACC patients. This protocol was implemented in January 2021 and continued until August 2022, when a randomized controlled trial (RCT) was initiated in our department. Patients with incomplete records, whose treatment plans were with sub-optimal placement of the applicators, whose plans were with sub-optimal dose prescriptions and did not follow standard loading pattern or deviated from the ICRU-38 guidelines due to a learning curve, were not eligible for analysis. 41 patients who met the eligibility criteria were included for analyses, all of whom received treatment under standard institutional protocol which began with EBRT in a dose of 45-50.4 Gy @ 1.8- 2 Gy per fraction, to the whole pelvis with concurrent weekly cisplatin. They were assessed to be fit for ICB within two weeks of completing EBRT to receive HDR ICB in 3-4 sessions of 7Gy or 2 sessions of 9Gy each. Both TR and tandem-ovoid (TO) applicators were used interchangeably for ICB sessions as successfully delivering the prescribed dosage to point A, while maintaining doses to critical organs within their permissible range could be achieved using both applicators equally. The standard institutional protocol ensured that the largest size TO or TR applicator, that a particular patient’s anatomy could accommodate, was utilized during every ICB session. First ICB session was performed utilizing either TO or TR applicator as per logistic availability and physician preference. Traditional vaginal packing (VP) technique was used for rectum separation alongside TO applicators and either conventional technique of using RR blade alone or indigenously modified technique to combine RR with customized VP was used alongside TR applicators for rectum separation. TR applicator with particular physical parameters (ring diameter, tandem length, tandem angle),when utilized during a ICB session on a particular patient alongside either conventional (RR alone) or modified technique (combination of RR and customized packing) for rectum separation, it was mandatory under standard institutional protocol to repeat TR applicator with same parameters during at least one of the subsequent sessions with alternate technique on the same patient. So that two ICB sessions done on one patient utilizing TR applicators differed only with regard to rectum separation technique. Rest of the sessions if applicable, were performed with the applicator type and technique which produced best dosimetric parameters during previous sessions. The authors who performed ICB procedures, were well-versed in both conventional and modified techniques as both techniques for separating the rectum are routinely employed during sessions utilizing TR applicators within our institution. The study protocol received approval from the Institutional Ethics Committee.

### Brachytherapy procedure

All sessions were carried out in the dedicated operation room for brachytherapy in our department under intravenous sedation and analgesia. After negotiating through the cervical canal, total length of uterine cavity from external os till fundus was measured with uterine sound and central tandem of measured length was inserted. A ring applicator of the largest diameter that could be accommodated in the upper vagina was then selected and threaded over the central tandem. The ring was positioned to encircle the cervix and interlocked with central tandem. During conventional procedures, standard RR blade was inserted and clamped with central tandem. Saline soaked gauze packing in the centre to stabilize the applicator assembly and in the anterior to displace bladder was then accomplished. But during modified technique, an additional step was that before the placement of RR blade, available space posterior and superior to the ring cap in the fornices and upper vagina was assessed to determine whether placement of additional vaginal packing would be possible. Radio-opaque VP was then carefully placed in layers, posterior to the ring cap in the fornix and upper vagina to enable additional rectum separation. The radio-opaque VP used is thin ribbon-shaped gauze in a shape of a roll, approximately 2 cm in width soaked in ixohexol contrast diluted with normal saline in a ratio of 1:1. Physician’s skill and judgement is required to customize VP for a range of vaginal spaces so that the space behind the ring cap, in the fornices and upper vagina, was optimally filled without any obstruction and undue resistance or pressure to the placement and assembly of the RR. If needed, some of the layers of gauze packing could be pulled back to allow for adequate space for accommodating RR blade, which is a crucial decision to avoid mucosal tear. After clamping the RR blade, packing in the centre to stabilize the applicator assembly and in the anterior to displace bladder is then accomplished in all procedures. Whole assembly is now kept softly pushed against the cervix to evaluate the remaining space between RR and lower part of posterior vaginal wall. If any scope for additional packing was identified, then further packing in the lower vagina, was carried out without disturbing the assembly, with the help of a long forceps, taking an approach from posterior aspect of the RR blade. The orthogonal radiographs were taken on conventional simulator (Varian, Acuity, USA) and the images were transferred to the treatment planning system (TPS) via Digital Imaging and Communications in Medicine (DICOM) connection.

### Brachytherapy treatment planning

The ICB treatment planning was done on the Oncentra Brachy v4.6 TPS and was delivered by Nucletron, Microselectron HDR V3 (18-channel) remote after-loading machine using Ir-192. The orthogonal images with applicators in place were reconstructed, and dwell positions were activated to recapitulate the familiar and symmetric pear shaped dose distribution typically used in low dose rate (LDR) ICB. We consistently practice a uniform loading pattern in our department (Figure 3). Point A was taken as 2cm superior to the lowermost intra-uterine source along the axis of intrauterine tandem and 2cm lateral in the same plane. The point B was taken as 2cm superior along the axis of intrauterine tandem and 5cm lateral in the same plane. The ICRU rectal point and bladder point were taken according to the recommendations of ICRU-38.^14^ The doses (both absolute and percentage) to these points were reported for each ICB session. Classical pear shaped reference volume (Vol_ref_) was measured and reported for each ICB session. The 60Gy reference volume (Vol_60Gy_), was measured and reported at the end when treatment of a particular patient was completed as the dose contributions from whole pelvic irradiation and all ICB sessions were to be taken into consideration. The total reference air kerma (TRAK) values for each fraction were also recorded.

**Figure 3.**
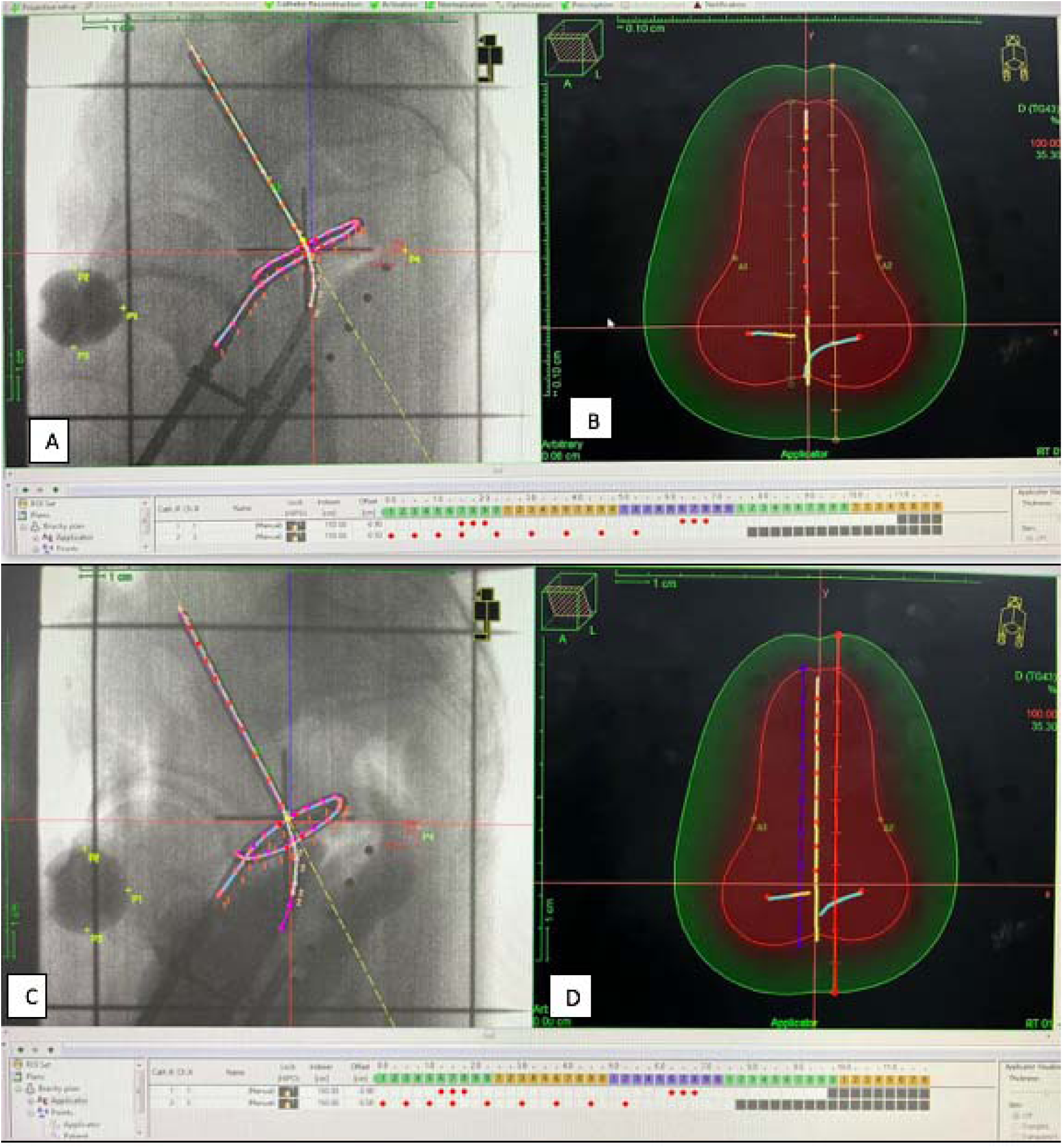
Treatment Planning images : A) Shows the reconstruction of the tandem-ring applicator with active dwell positions in red dots. Dwell position activation (loading pattern) can be seen at the bottom of the image A & B with red dots corresponding to respective dwell positions for both ring and tandem. The image also shows the ICRU bladder (P1) and rectum (P4) points. (B) Shows the classical pear shape reference volume shaded in the red and 60 Gy volume in this patient is represented by 35.3 % isodose curve line shaded in green for this ICB session performed with conventional technique. C) Shows the reconstruction of the tandem-ring applicator during ICB session performed with modified technique. Active dwell positions shown in red dots. Dwell position activation (loading pattern) can also be seen at the bottom of the image C & D with red dots. D) Shows a pear shape reference volume shaded in the red and 60 Gy volume is shown by 35.3 % isodose curve line shaded in green for this session with modified technique. When the same dose 7Gy (100%) was prescribed to point A, ICRU rectal point dose was 51.29 % (3.59 Gy) with conventional versus 35.31% (2.47 Gy) with modified technique. There was 16% (1.12Gy) reduction in the ICRU rectal point dose in this patient. The V ref (d_h_ X d_w_ X d_t_) i.e. the volume encompassed within 100% isodose curve, was similar at 177.94 cc with conventional versus 175.66 cc with modified technique. The V 60Gy (D_H_ X D_W_ X D_T_) was also similar at 767.14 cc with conventional versus 710.74 cc with modified technique. The dose to point B was similar at 26.9% versus 26.7% and ICRU bladder point dose was also similar at 28.86% (2.02 Gy) with conventional versus 23.58% (1.65 Gy) with modified technique. TRAK values were also similar at 0.00457 Gy versus 0.00461 Gy. ICRU: International Commission on Radiation Units and measurements; P 1: ICRU bladder point; P 4: ICRU rectal point; TRAK: Total reference Air Kerma; V ref: Reference volume; V 60 Gy : ICRU 60 Gy reference volume. TPS: treatment planning system

### Statistical analysis

The data was analysed using Stata Software version 15. Qualitative data was presented as frequencies and their percentages while quantitative data was described as means and their standard deviations. Student t test was used to compare the means of outcomes between two techniques. During the subset analysis a paired sample t test was used to compare means while analyzing outcome among two techniques performed on same patients. A two sided p value of less than 0.05 was considered as statistically significant.

## Results

A cohort of 41 patients received HDR ICB after completing whole pelvis EBRT, either to a dose of 46Gy/23 fractions (in thirty patients) or 50Gy/25 fractions (in eleven patients). Out of 41 patients, 20 received four sessions of 7Gy each, 14 received three sessions of 7Gy each, four received two sessions of 9Gy each, one received three sessions of 7Gy each and fourth session of 4Gy, one received two sessions of 7Gy each and third one of 4Gy, and one patient received four sessions of 4Gy each. This yielded a total of 141 ICB sessions. The reason for lowering the prescription dose to 4Gy during six sessions on 3 patients was to keep the organs at risk doses within their tolerance limit. The dose was prescribed to Point A and there was one-week interval between two sessions of brachytherapy in all the patients. The LDR equivalent median dose of 81Gy (range,70-90, mean 81.5 Gy) to Point A could be delivered, considering the contribution from both EBRT and all sessions of HDR ICB, while respecting ICRU rectal point tolerance limits of 75Gy or less and bladder point limit of 80Gy or less. Out of total 141 ICB sessions, 111 performed using TR applicators were analyzed in this study. Among these 111 ICB sessions, 44 in which RR blade alone was used constitutes conventional group and 67 in which a combination of radio-opaque VP and RR blade was used for rectum separation constitutes modified group.

### Patient characteristics

Baseline patient, stage of the disease and treatment related characteristics are shown in Table 1.

**Table 1.**
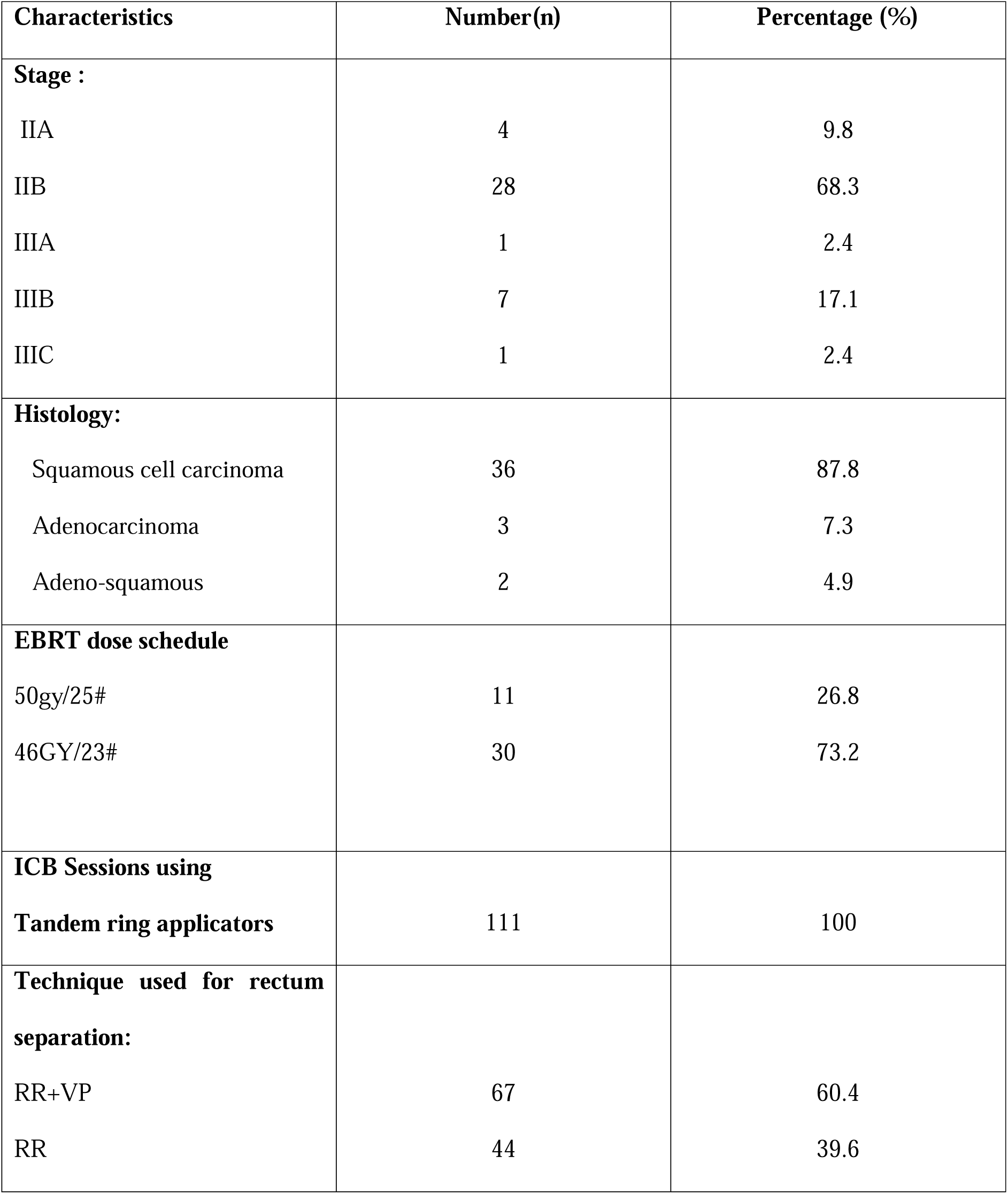
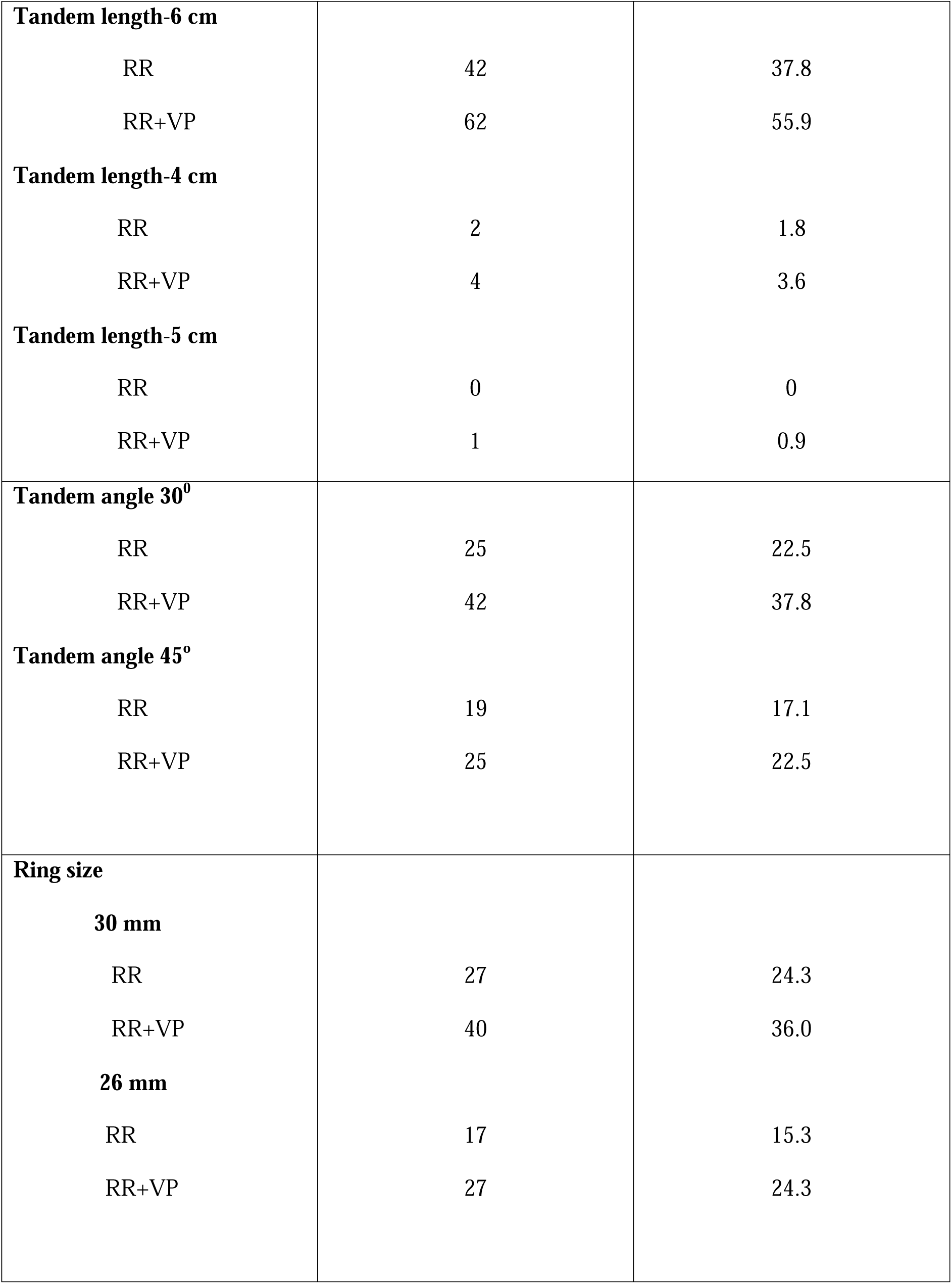
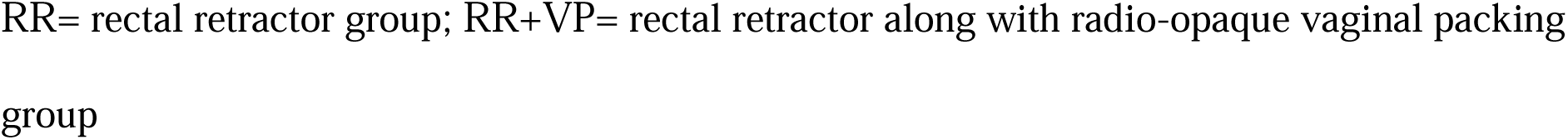
Distribution of stage- and applicator-related characteristics in the two groups.

| Characteristics | Number(n) | Percentage (%) |
| --- | --- | --- |
| **Stage :** |  |  |
| IIA | 4 | 9.8 |
| IIB | 28 | 68.3 |
| IIIA | 1 | 2.4 |
| IIIB | 7 | 17.1 |
| IIIC | 1 | 2.4 |
| **Histology:** |  |  |
| Squamous cell carcinoma | 36 | 87.8 |
| Adenocarcinoma | 3 | 7.3 |
| Adeno-squamous | 2 | 4.9 |
| **EBRT dose schedule** |  |  |
| 50gy/25# | 11 | 26.8 |
| 46GY/23# | 30 | 73.2 |
| **ICB Sessions using**<br>**Tandem ring applicators** | 111 | 100 |
| **Technique used for rectum**<br>**separation:** |  |  |
| RR+VP | 67 | 60.4 |
| RR | 44 | 39.6 |
| **Tandem length-6 cm** |  |  |
| RR | 42 | 37.8 |
| RR+VP | 62 | 55.9 |
| **Tandem length-4 cm** |  |  |
| RR | 2 | 1.8 |
| RR+VP | 4 | 3.6 |
| **Tandem length-5 cm** |  |  |
| RR | 0 | 0 |
| RR+VP | 1 | 0.9 |
| **Tandem angle 30°** |  |  |
| RR | 25 | 22.5 |
| RR+VP | 42 | 37.8 |
| **Tandem angle 45°** |  |  |
| RR | 19 | 17.1 |
| RR+VP | 25 | 22.5 |
| **Ring size** |  |  |
| **30 mm** |  |  |
| RR | 27 | 24.3 |
| RR+VP | 40 | 36.0 |
| **26 mm** |  |  |
| RR | 17 | 15.3 |
| RR+VP | 27 | 24.3 |

RR= rectal retractor group; RR+VP= rectal retractor along with radio-opaque vaginal packing group

### Dosimetric parameters

The comparison between dose and volume parameters is shown in Table 2. For the same dose prescribed to point A in both groups, the mean dose to the ICRU rectal point was 44.1% of the prescription dose (range, 23.8-77.8%; median 42.5%) in the modified group and 55.5% (range, 36.4-73.1%; median 56.5%) in the conventional group. There was 11.4% reduction in the mean dose to the ICRU rectal point (p<0.001) in the modified group compared to the conventional group. The mean dose to the ICRU bladder point was 55.5% of the prescription dose (range, 14.8-127.2%; median 55.5%) in the modified group and 49.8% (range 11.6-95.6%; median 51.5%) in the conventional group. There was 5.7% increase in the mean dose to the ICRU bladder point (p=0.21). The mean dose to point B was similar at 25.7% of the prescription dose (range, 22.2-27.2%; median 26.0%) in the modified group, and 25.9% (range, 23.3-27.7%; median 26.1%) in conventional group. The mean reference volume (Vol_ref_) for the brachytherapy session expressed as d_h ×_ d_w_ ×d_t,_ the mean 60Gy reference volume (Vol_60Gy_) expressed as H×W×T and the mean TRAK value (Gy/hr at 1 m) were similar between the two groups.

**Table 2.** Comparison of dose and volume parameters.

| Physical parameter | N (fractions) | Mean | SD | t value** | P value* |
| --- | --- | --- | --- | --- | --- |
| **Point B dose**<br>(% of prescription) |  |  |  |  |  |
| RR | 44 | 25.9 | 1.04 | 0.95 | 0.34 |
| RR+VP | 67 | 25.7 | 1.09 |  |  |
| **ICRU rectal point dose**<br>(% of prescription) |  |  |  |  |  |
| RR | 44 | 55.5 | 7.9 | 5.86 | *<0.001* |
| RR+VP | 67 | 44.1 | 11.2 |  |  |
| **ICRU bladder point dose**<br>(% of prescription) |  |  |  |  |  |
| RR | 44 | 49.8 | 20.1 |  |  |
| RR+VP | 67 | 55.4 | 25.3 | -1.26 | 0.21 |
| **ICRU reference volume**<br>(V<sub>ref</sub>)(d<sub>h</sub>×d<sub>w</sub>×d<sub>t</sub>) in cc |  |  |  |  |  |
| RR | 44 | 174.9 | 12.2 | 1.22 | 0.20 |
| RR+VP | 67 | 171.6 | 14.8 |  |  |
| **ICRU 60 Gy volume**<br>(V<sub>60Gy</sub>) (H×W×T) in cc |  |  |  |  |  |
| RR | 44 | 703.9 | 199.6 | -0.36 | 0.71 |
| RR+VP | 67 | 717.6 | 189.0 |  |  |
| **TRAK value** |  |  |  |  |  |
| RR | 44 | 0.0046 | 0.0005 | 1.63 | 0.10 |
| RR+VP | 67 | 0.0044 | 0.0007 |  |  |

RR= Conventional group; RR+VP= Modified group; ICRU= International Commission on
Radiation Units & Measurements; TRAK: total reference air kerma
SD= standard deviation. \* Statistically significant p values are shown in bold italic print.
\*\* A student t test was used for comparison between two groups.

In this detailed paper we found it appropriate to add a subset paired analysis as two groups were not matched as numbers of sessions in modified group were 67 and in conventional group 44. This is largely because the protocol allowed the last session of ICB to be done with the technique that yielded best dosimetric parameters during the previous sessions in that particular patient. Also in two patients all ICB sessions were done with modified technique inadvertently and for the same reason in one patient all sessions were found to be done with conventional technique, despite the departmental protocol which mandated that the subsequent session was to be done with the alternate technique using same applicator. Moreover, in one patient all sessions were done with conventional technique, as during first session with conventional technique while placing RR blade, small mucosal tear occurred which was managed conservatively and modified technique was not attempted during subsequent sessions. We omitted these four patients from the subset analysis due to their consistent utilization of a singular technique in all sessions, which hindered our ability to carry out an intra-patient comparison of two techniques. This exclusion served to eliminate any potential bias in selection.

To perform paired subset analysis on the remaining 37 patients, we analyzed two sessions per patient that incorporated consistent utilization of identical physical parameters of the applicator, such as ring size, tandem length, angle, and loading pattern (Table 3).

**Table 3.**
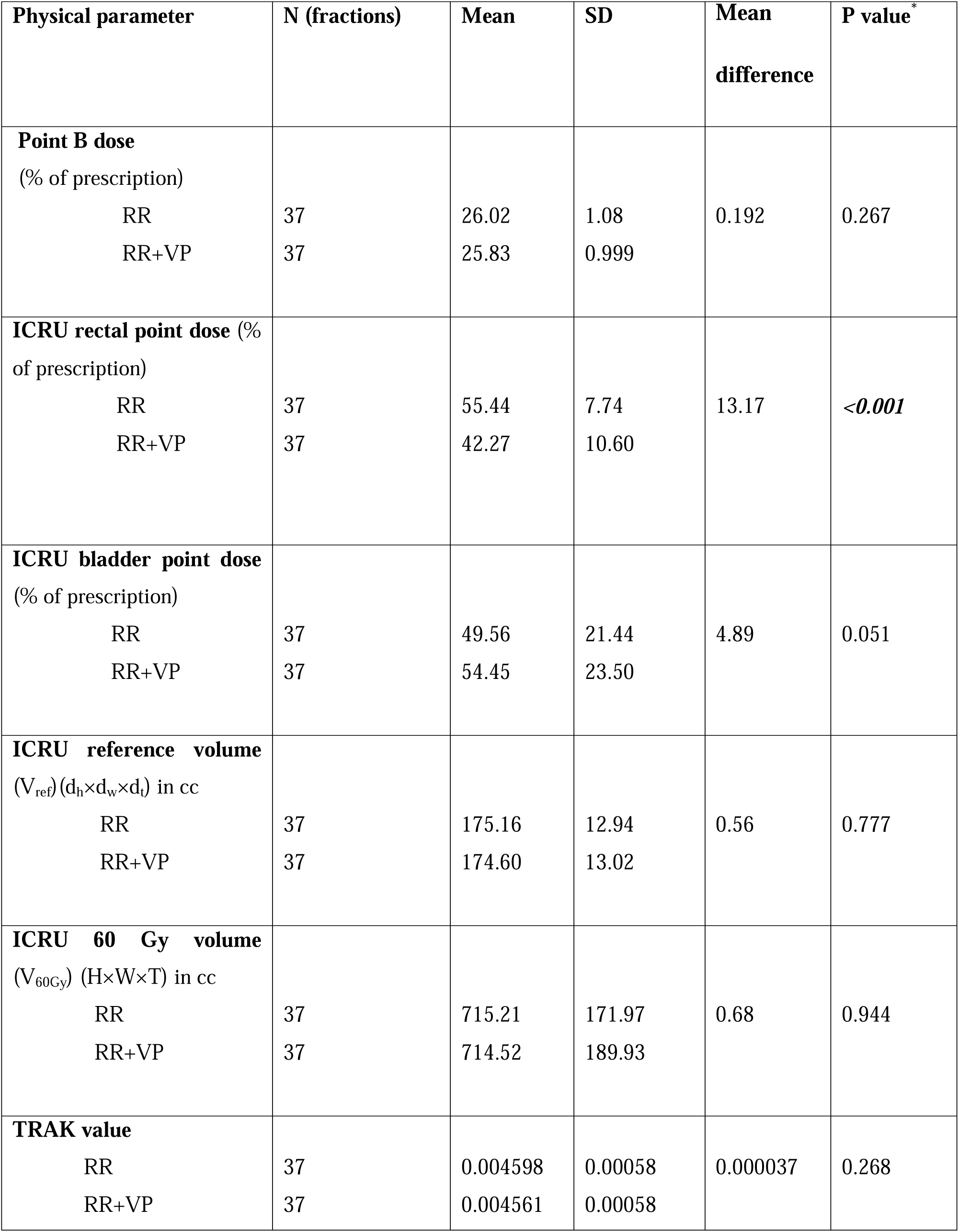
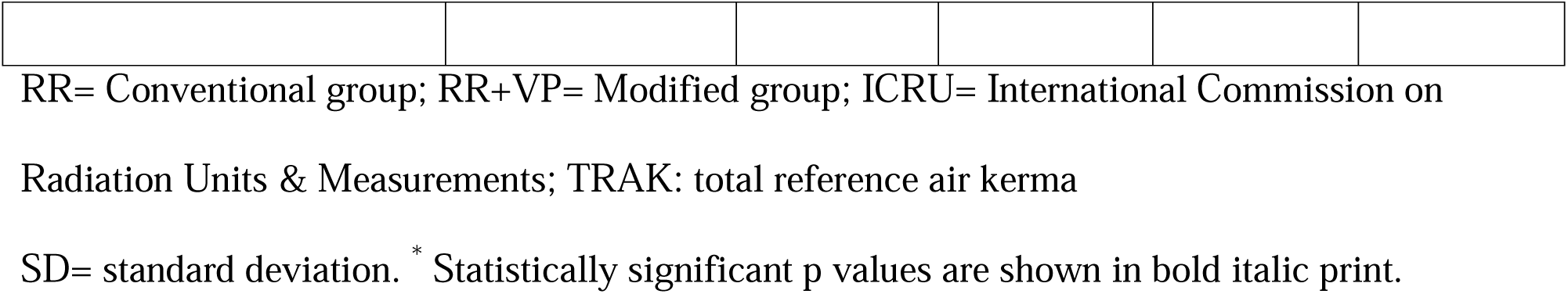
Paired Subset analysis among two treatment groups.

For the same dose prescribed to point A in both groups, there was 13.17% reduction in the mean dose to the ICRU rectal point (p<0.001) in the modified group compared to conventional group. There was 4.89% increase in the mean dose to the ICRU bladder point (p=0.051) in modified group compared to conventional group. All other dosimetric parameters had no statistically significant difference between two groups.

## Discussion

The modified technique, which involves combining VP with the RR blade, resulted in an average reduction of 11.4% (p<0.001) in the dose to the ICRU rectal point (D_ICRU_ for rectum) per ICB session compared to the conventional approach of using the RR blade alone for rectum separation. In the paired subset analysis, the average reduction in D_ICRU_ for rectum was 13.17% (p<0.001). This technique was applied to the majority of women (39 out of 41) whose anatomy allowed for the placement of ring applicators with the RR blade. In one patient the fornices were so shallow that even inserting the RR blade was challenging. Therefore, the modified technique may be used on the patients whose anatomy allows the placement of ring applicators with RR blade without over stretching the mucosa and not on the patients whose anatomy doesn’t allow their insertion and placement.

In this study, there was an increase of 5.7% (p=0.21) in the mean dose to the ICRU bladder point (D_ICRU_ for bladder) per session. In the paired subset analysis, the average increase in D_ICRU_ for bladder was 4.89% (p=0.051). This simultaneous increase in the mean dose to the ICRU bladder point was 10.1% (p=0.024) in our previous study using this technique.^6^ Since then, we have started paying close attention to the anterior packing as well. This study also demonstrates that by giving deliberate consideration to the anterior packing, we could minimize the simultaneous increase in the bladder point dose associated with this technique.

The customized VP is placed in the posterior fornices (behind the ring) and the upper vagina to a limited extent, allowing the RR blade to be positioned and assembled without any obstructions. The aim is to supplement the rectal retraction achieved primarily with the standard RR blade, with the combination of customized VP.

The dose was prescribed to point A and a uniform loading pattern was followed. The doses to organs at risk (OARs) were recorded and reported as codified in the ICRU-38 report.^14^ It is still relevant as orthogonal X-rays and CT scans are the primary imaging modalities for treatment planning in low and middle-income countries (LMICs), which contribute to a majority of the cervical cancer burden. In LMICs, MRI being utilized in only 5-9% of cases and also majority of the centres are prescribing the dose to point A. Even during transition from 2-D to 3- D based parameters, point A based prescription is preferred.^15,16^ Moreover, despite 3-D image guided adaptive brachytherapy (IGABT) using magnetic resonance imaging (MRI) is being advocated as gold standard, the efforts to adopt MRI-based planning in all patients were associated with unique implementation difficulties raising the concern for inferior local control due to increased overall treatment time. In a recent study representing the real-world scenario in LMICs, authors suggested a rational implementation of point-based brachytherapy even in this era of MRI-guided adaptive brachytherapy in patients with good response and limited residual disease at the time of ICB, to optimize the utilization of available resources for achieving best possible clinical outcome.^17^

In centres employing orthogonal x-ray-based planning, doses are recorded and reported for the ICRU rectum and bladder reference points to represent the doses to these OARs. In centres utilizing volumetric imaging, the OARs (bladder and rectum) are delineated in axial sections, and doses to 2cc volumes are reported.^16^ Significant increase in grades of rectal toxicity in parallel to an increase in mean doses to both D_2cc_ (minimum dose to most exposed 2cc volume of rectum) and D_ICRU_ (dose to ICRU rectal point) is well established.^10–12^ A vast experience gained in the past with the ICRU rectal point which has been used as a reference for decades, still holds good as a high correlation has been shown between the average values of D_2cc_ and D_ICRU_ and the dose level of the D_2cc_ found to be closer to the D_ICRU_ for rectum.^10,18–21^ Therefore, modified technique may also be evaluated by the providers who are practicing 3-dimentional (3D) IGABT, delineating low, intermediate and high risk clinical target volume (CTV) for dose prescription and contouring OARs for reporting D_2cc_, D_1cc_, D_0.1cc_ (doses to most exposed 2cm^3^, 1cm^3^, 0.1cm^3^ of respective OAR), as per The Groupe Europeen de Curietherapie (GEC)-ESTRO working group recommendations. It has been noted in previous studies that the implementation of strategies for organ retraction and ongoing enhancements in applicator design can yield added advantages in achieving improved dosimetric distributions.^22–24^

The study highlights that the integration of personalized vaginal packing with the rectal retractor blade exceeds the effectiveness of solely relying on the rectal retractor blade for achieving rectum separation. This is especially important in light of the current trend of expanding the use of TR applicators with RR blade for rectum separation to include all patients undergoing ICB, rather than limiting their use to just the ideal subset.

The limitation of the study is that it is an observational study in which 3 to 4 HDR ICB sessions were conducted on each patient in accordance with a predetermined departmental protocol.

Though the protocol stipulated that after using a specific technique for rectum separation with the TR applicator in one session, a different technique with the tandem-ring applicator, having identical physical parameters, should be employed in subsequent sessions for each patient. These subsequent sessions were not necessarily consecutive; logistical constraints in a high-volume center with limited resources permitted the use of TO applicators in one or two out of the total 3 to 4 HDR ICB sessions per patient. Therefore, there could be a two to three weeks interval between two comparable sessions. As decades of experience suggests that during the course of EBRT followed by ICB, there is progressive regression of tumour with associated anatomical changes, an ideal study would randomize first session utilizing a particular applicator and rectum separation technique and consecutive second session with same applicator parameters but alternate rectum separation technique to negate the possible confounding effect of variation in anatomy even in the same patient during the course of treatment. Moreover, despite the implementation of a consistent loading pattern in our department, there existed variability among physicists primarily attributable to the learning curve, prior to attaining a standardized loading pattern. It is only in the subset analysis that all the planning parameters are nearly perfectly aligned. Another limitation is that the results of the study are with X-ray based planning as per ICRU-38 recommendations where limited optimization of dwell positions and times is allowed and not yet validated in 3-D image guided brachytherapy (IGBT).

## Conclusion

The mean radiation dose delivered to the ICRU rectal point with the conventional use of tandem- ring applicators and a standard rectal retractor blade is significantly reduced by addition of a customized vaginal packing with the help of a novel packing technique in patients undergoing intracavitary brachytherapy for locally advanced cervical cancer. The suggested technique to combine standard rectal retractor blade with customized vaginal gauze packing is easily applicable, cost-effective, and therefore can be embraced by providers worldwide, particularly in settings with limited resources. Utilizing this technique, the average decrease in the mean dosage to the ICRU rectal point (expressed as a percentage of point A dose) is 11.4% per session compared to the traditional method of solely using a rectal retractor (p*<0.001*). In the subset analysis, the average reduction in rectal point dosage per session is 13.17% (p<0.001).

## Data Availability

All data produced are available online at Harvard Dataverse Dataset Replication Data for A retrospective analysis to compare the dosimetric parameters of two rectum sparing techniques during intracavitary brachytherapy with tandem–ring applicators for cervical cancer

https://doi.org/10.7910/DVN/13BFHP

## Acknowledgments

This study is dedicated to our teacher and mentor Professor Rajeev Kumar Seam. The authors acknowledge Professor Anupam Parashar for providing conducive environment for research in our institution.

